# Pulmonary arteriovenous malformation risks: Single-centre hybrid study of 1,149 cases re-emphasises hypoxemia without pulmonary hypertension, and paradoxical embolic/infective stroke etiologies

**DOI:** 10.64898/2026.05.07.26352680

**Authors:** Claire L Shovlin, Nicola M Coote, Rhea Sheth, Hadrien Janbon, Misha Iyer, Hannah C Tighe, Heidi McKernan, James Springett, Brendan Mallia Milanes, Nicola Read, Joe An Cabantug, Julie Ranger, Hemanth Prabhudev, May Al Sahaf, Ben Glampson, Erik Mayer, Ali Alsafi

## Abstract

Pulmonary arteriovenous malformations (PAVMs) larger than 4mm in size are estimated to affect 38 per 100,000 individuals [95% confidence intervals 18-76]. They provide an anatomical right-to-left shunt such that each heartbeat, a proportion of the cardiac output bypasses the pulmonary capillary bed, preventing essential processing functions such as gas exchange and filtration of blood-borne emboli. Although large cohort series were published in earlier decades, more recent data series have been scant. Here we report features of 1149 consecutive patients with imaging-proven PAVMs, reviewed at a single UK centre between 1984-2026, including 813 [95% confidence intervals 0.68, 0.74] with clinical and/or genetically confirmed hereditary hemorrhagic telangiectasia (HHT). The median age was 47y, and 735 [0.64, 0.48] were female. We report 4348 oxygen saturation measurements at presentation and follow-up, and 810 pulmonary artery pressure (PAP) measurements made at angiography prior to treatment of PAVMs by embolisation. Together, these confirm that there is no risk of hypoxic pulmonary hypertension, with PAP measurements higher in patients with higher SaO_2_. Massive hemoptysis or hemothorax affected 18/1,149 [0.007, 0.018] patients, confirmed ischemic strokes 125/1,149 [0.09, 0.13] patients, brain abscess 107/1,149 [0.08, 0.11] patients, and hemorrhagic strokes 23/1149 [0.01, 0.03] patients. These data will inform design of future work to evaluate etiologies, associations and implications for clinical practice.

## BACKGROUND

Pulmonary arteriovenous malformations (PAVMs) provide an anatomical right-to-left shunt which commonly causes hypoxemia due to impaired gas exchange.^1^ Clinically, PAVMs are an important cause of strokes due to blood-borne particulate matter crossing from the venous to arterial circulation, instead of being filtered out by pulmonary capillaries.^2^ PAVMs cause ischemic stroke where they are responsible for a substantial proportion of stroke-in-young,^3^ and brain abscess, an otherwise rare condition in immunocompetent individuals.^4^ Prevalence data showed PAVMs of 4mm diameter or larger affected 38 per 100,000 people [95% confidence intervals 18, 76],^5^ although smaller PAVMs are increasingly detectable by conventional thoracic computer tomography (CT) scans. Most PAVMs occur in association with hereditary hemorrhagic telangiectasia (HHT) which is inherited as an autosomal dominant trait.^6^ Major HHT manifestations including AVMs in the lungs, liver and brain; telangiectasia in the nasal and gastrointestinal tracts that commonly bleed; and hepatic AVM-associated post-capillary pulmonary hypertension.^6^

Pre 2018 data series containing up to 445 consecutive patients were used to inform major clinical statements on management.^2,7,8^ Our goal was to provide updated information from a more modern series.

## METHODS

With ethical approval from the South West-Central Bristol Research Ethics Committee (21/SW/0120), the Imperial iCARE database holds deidentified electronic patient record data of patients reviewed at Imperial College Healthcare NHS Trust. A Rare Disease database was constructed to hold the routine data from National Health Service (NHS) clinical care for all patients with known PAVMs reviewed in the Imperial/Hammersmith pediatric and adult PAVM services from 29/05/1984. Data were deidentified by the NIHR Imperial Biomedical Research Centre (BRC) iCARE team before hosting in the iCARE Secure Data Environment. Project approval of the project “Strokes and pulmonary arteriovenous malformations” was by the January 2026 NIHR Imperial BRC Data Access and Prioritisation Committee Meeting.

For the iCARE database, retrospective lists of patients seen only prior to 1999, or solely in pediatric or embolisation services were added to a prospective database of patients seen in adult medical PAVM services at Hammersmith Hospital from 1 May 1999-09 April 2026 which had been generated with ethical approval from the Hammersmith and Queen Charlotte’s & Chelsea Research Ethics Committee (REC ref. 2000/5764). Primary digitised, electronic and pre-2019 paper records were reviewed to capture, validate and date record measurements across all service reviews, including metrics not examined in any previous research database.

Briefly, PAVMs were diagnosed by thoracic computer tomography (CT) scans, except for some very early patients who progressed to angiography following chest x-ray and functional shunt assessments. For functional quantification of right-to-left shunting that had been validated against same-day ^99m^technetium-labelled albumin macroaggregate perfusion scans,^1^ the oxygen saturation (SaO_2_) was measured by pulse oximetry for 10 minutes standing (erect), with the mean values of minutes 7-10 recorded.^9^ Pulmonary artery pressure (PAP) was measured at angiography immediately before embolisation where indicated: PAP measurements were not made in 12 patients with pre-existing pulmonary hypertension where angiography with a view to embolisation was not offered due to the risks of precipitating a fatal increase in PAP,^8,10,11^ or embolisation was not considered appropriate given the small size of PAVMs.^7^ HHT was diagnosed clinically by 3 or more Curaçao criteria,^12^ or an NHS genetic diagnosis (a clinical gene test report of a heterozygous pathogenic or likely pathogenic variant in *ENG, ACVRL1, SMAD4* or *GDF2*).^6^ Symptoms, comorbidities and treatments were recorded, alongside all clinically-defined strokes lasting >24 hours with etiologies as defined by NHS reports and treatments. There were separate categories for strokes of unknown, traumatic or iatrogenic etiology. Events where stroke etiology was not confirmed with clinical differentials including migraine and seizures were recorded as ‘possible stroke’.

To capture other recent cohort studies, a PubMed search was conducted on 30/04/2026 for the term “pulmonary arteriovenous”, limiting to records published after the British Thoracic Society Clinical Statement in December 2017^2^. Titles and abstracts were used to assign publications to categories of ‘not relevant’ and ‘relevant’. ‘Not relevant’ included PAVMs induced by cavopulmonary shunt surgery for cyanotic congenital heart disease, since there were no post-surgical cases in the current series. ‘Relevant’ publications were sub-categorised into case reports, reviews/editorials, corrections/replies, and primary data papers. Primary data papers were sub-categorised into embolisation, surgery and medical categories, with medical category publications reviewed in detail.

## RESULTS

### General demographics

By 09/04/2026, the database contained data on 1149 patients with radiologically confirmed PAVMs (Figure 1a). 735/1149 [0.64, 0.48] were female. The median age at first review was 47y (interquartile range [IQR] 33, 61y), and at last review 52y [IQR 39, 64y], providing 4724 years of follow-up. Clinically and/or genetically diagnosed HHT was confirmed in 813/1,149 [0.68, 0.74] (Figure 1b). The non-HHT subcohort was more female-rich (236/336 [0.653, 0.75], than the HHT subcohort (499/813 [0.58, 0.647, p=0.0044 (Figure 1c).

**Figure 1:**
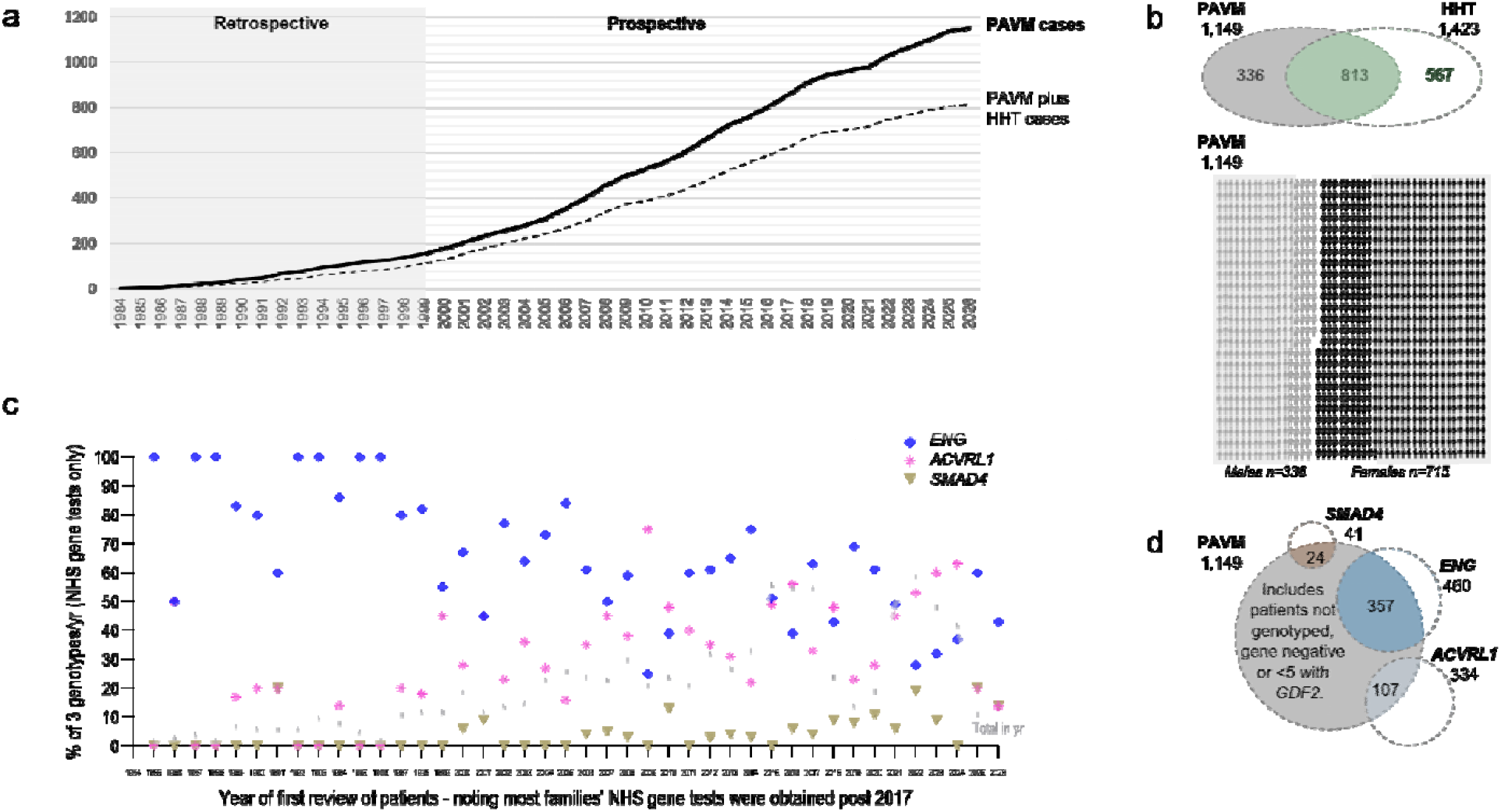
Cohort of 1,149 patients with radiologically confirmed PAVMs. **a)** Cumulative rates by year of first presentation. **b)** Overview of proportion of patients with PAVMs (grey), HHT (green) and both diagnoses: Upper panel all patients, lower panel, patients categorised by sex. **c/d)** Genotypes assigned to patients and their families by c) year of patient’s first presentation, and d) overall PAVM numbers. The smaller number of *GDF2* patients/families (n<5) are not shown.

In 483/813 [0.56, 0.63] patients with both radiologically-confirmed PAVMs and HHT, the family HHT genotype was identified through clinical gene tests. Genotypes changed between 1984-2026 (Figure 1c) reflecting a greater *ENG* bias in early years when patients were solely referred for PAVM care, and a broader later range of genotypes as patients were also referred for HHT management. By 09/04/2026, the PAVM cohort of *ENG* (n=357*), ACVRL1* (n=102*), SMAD4* (n=24) represented 82%, 32% and 62% of HHT patients with the respective genotype (Figure 1d).

### Physiological parameters by genotype

Earlier PAVM cohorts did not refer to HHT genotypes. We therefore evaluated whether metrics differed according to genotypes. At first presentation, hypoxemia, a biomarker for functional RL shunting was more severe in *ENG* patients (p<0.0001, Figure 2). Hemoglobin is increased as an adaptation to PAVM-induced hypoxemia and reduced if iron deficiency develops (in this cohort, usually due to HHT nose bleeds), but no trends were evident between the genotypes (Figure 2). In patients undergoing embolisation, PAP were marginally higher in *ACVRL1* patients (Figure 2). We concluded that there were some genotypic differences, but not sufficient to restrict analyses to the genotyped population.

**Figure 2:**
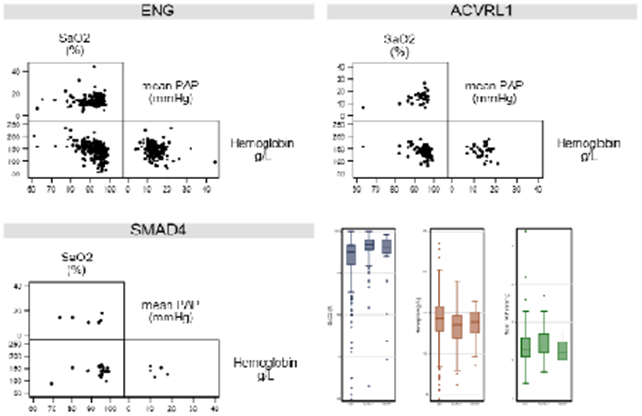
Demographics of 483 patients with radiologically confirmed PAVMs and a confirmed *ENG, ACVRL1*, or *SMAD4*. pathogenic or likely pathogenic variant identified for their family. Note the broad spread of values in the diagonal scatter plots by genotypes. Box plots are ordered *ENG, ACVRL1, SMAD4*. There are trends for patients with PAVMs associated with *ACVRL1* to have less severe PAVMs (higher SaO_2,_ navy), lower hemoglobin (red), and higher mean PAP (green) than the other genotypes. Boxes indicate medians and interquartile range, error bars indicate 95^th^ percentiles and outliers are indicated as circles.

### Physiological parameters by time-series

The largest previous series examining ischemic stroke risk had reported patient data up to February 2013.^9^ To examine whether PAVM demographics or complications had changed since that period, we sub-categorised the cohort of 1,149 cases into the 616 who were first seen before March 2013, and 533 who were first reviewed in/after March 2013. Figure 3a indicates that the earlier series had significantly lower pre-treatment SaO_2_ (p<0.0001). However, post-treatment SaO_2_ (Figure 3b), the inverse relationship with hemoglobin reflecting compensatory polycythemia (Figure 3c) and resulting maintenance of arterial oxygen content (CaO_2_, Figure 3d) were similar in both subcohorts.

**Figure 3:**
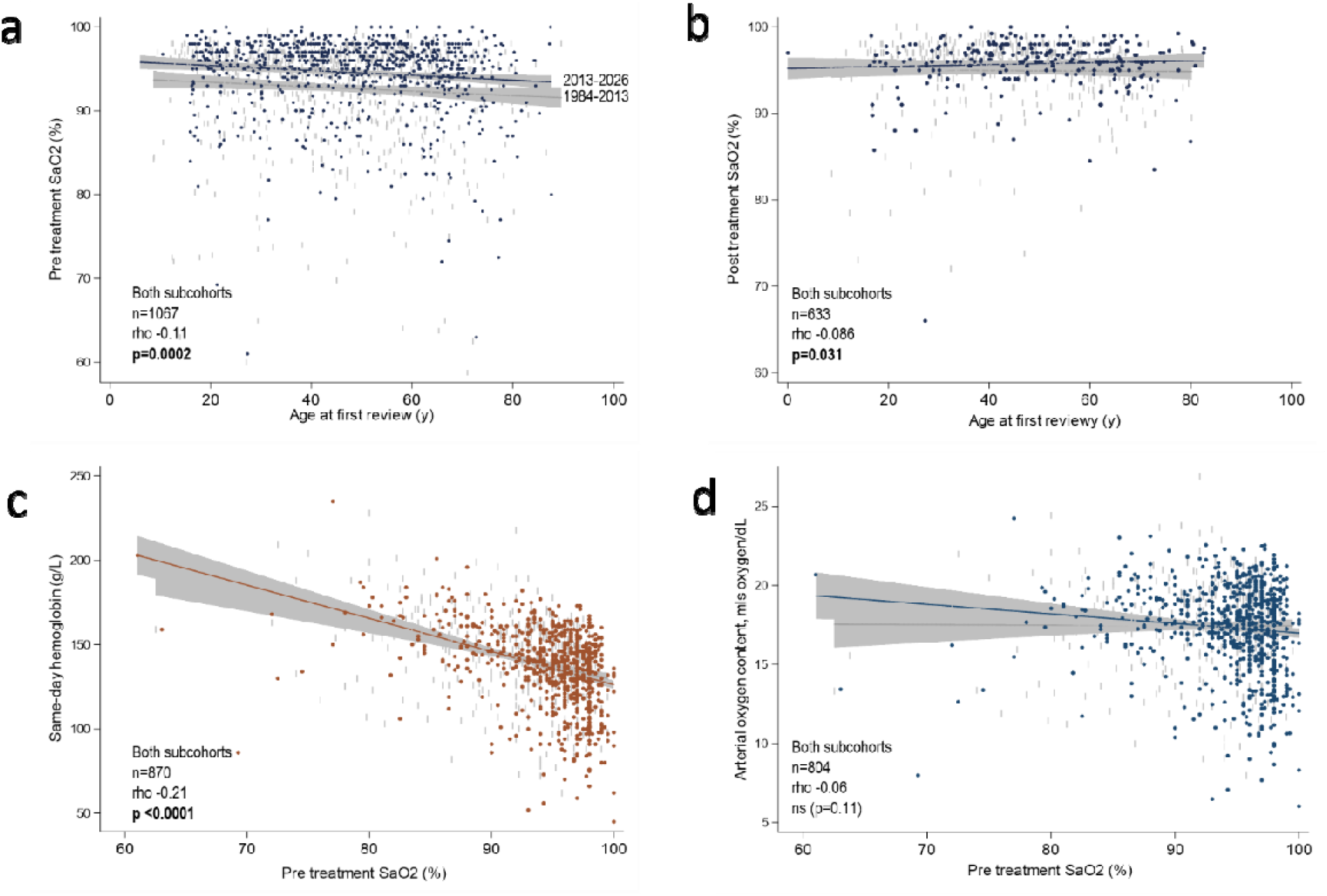
Oxygenation comparisons between patients presenting with PAVMs early (1984-Feb 2013) or later (March 2013-April 2026). Two-way plots with linear fit lines, 95% confidence intervals for subcohorts and Spearman rho with p-values for correlations across both subcohorts for: **a)** Mean pre-treatment SaO_2_ by age at first review; **b)** Mean post-treatment SaO_2_ by age; **c)** Same-day hemoglobin by SaO_2_; **d)** Arterial oxygen content, CaO_2_ calculated as 1.34*hemoglobin*same-day SaO_2_.

The 561 patients with PAP measurements had similar mean PAP pre-treatment to the 1,187 general population subjects whose data defined pulmonary hypertension (reference series 14.0±3.3 mmHg^13^; 1984-2013 PAVM subcohort 14.66 mmHg [SD 5.04]; 2013-2026 PAVM subcohort 14.03 mmHg [SD 4.47]). Overall, as shown in Figure 4a, >90% of pre-treatment PAP measurements were within the normal range (≤20mmHg^13^). In both cohorts, as in the general population, mean PAP increased with age, though the age-related trend was less pronounced in the more recent series (rho 0.23 vs 0.34, Figure 4a). This difference was maintained after adjusting for sex and SaO_2_ either at presentation, or at the time of PAP measurement. Contrasting to hypoxic pulmonary hypertension, lower SaO_2_ were not associated with higher PAP (Figure 4b). Instead, there was a trend for lower SaO_2_ to be associated with lower PAP (p=0.018, Figure 4b).

**Figure 4:**
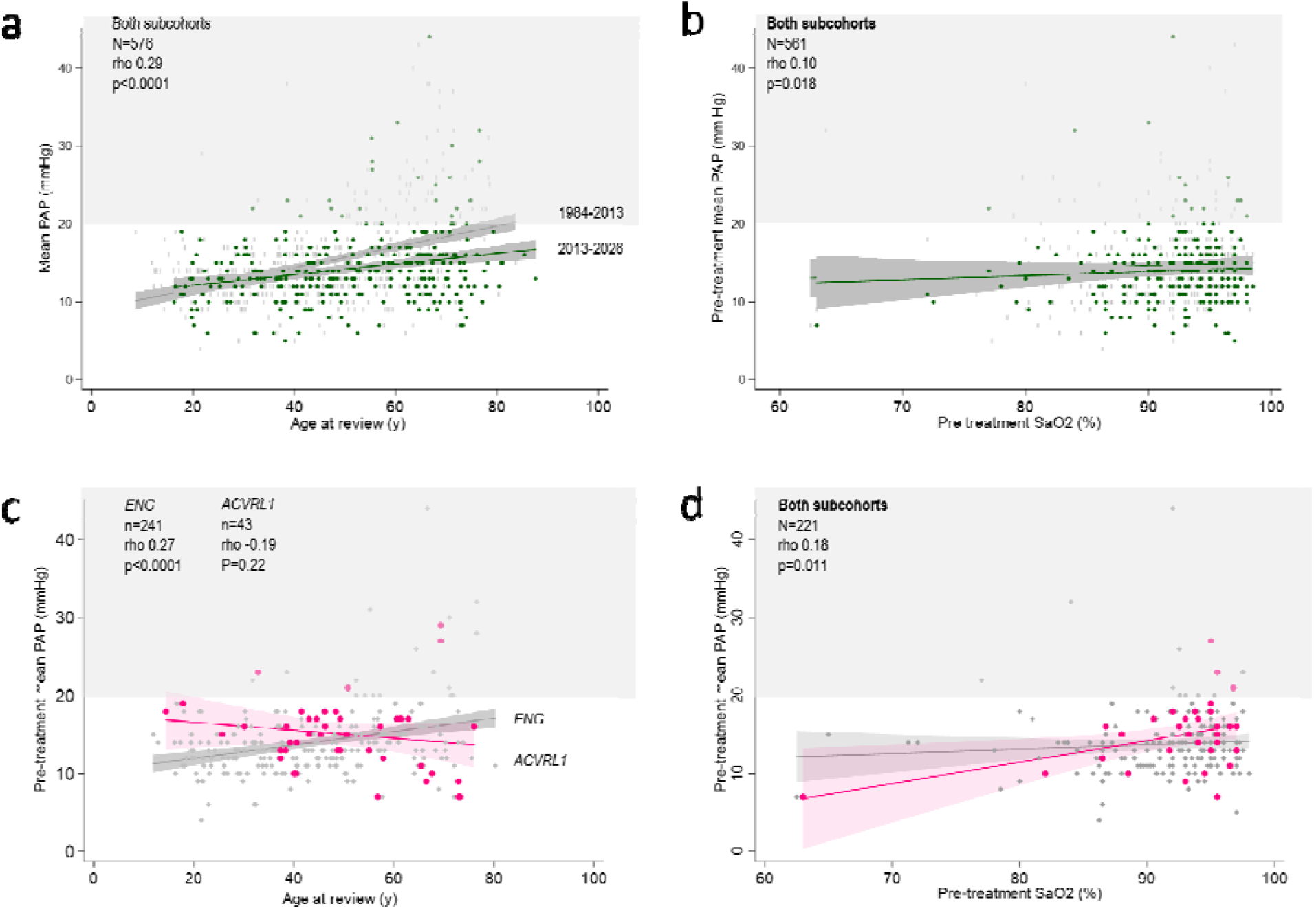
Pulmonary artery pressure comparisons for patients undergoing PAVM embolisation. Two-way plots with upper shaded range indicating pulmonary hypertension values post reference^31^ (previously, the upper limit or normal was 25mmHg). Linear fit lines and 95% confidence intervals as indicated for subcohorts with Spearman rho and p-values: **a/b)** Cohorts by first review 1984-2013, or 2013-2026: **a)** Serial mean PAP measurements by age at first review. Note crude PAP (first or last) did not differ between cohorts (Mann Whitney p values> 0.05). **b)** Pre-treatment mean PAP by pre-treatment SaO_2_, generally measured same-day or previous day: Note that if hypoxic pulmonary hypertension were induced by PAVM-derived hypoxemia, the fit line would have the opposite slope. Instead, there was a trend to an association between higher SaO_2_ and higher PAP. **c/d)** Cohorts by *ENG* or *ACVRL1* HHT genotype.

Given the higher risk of pulmonary arterial hypertension (PAH) with *ACVRL1* genotypes^6,14^ but relatively minor differences in the PAVM patient data (Figure 2), we compared the two major HHT genotype groups for PAP relationships with age and SaO_2_. There was no significant change with age for the *ACVRL1* subcohort but PAP measurements for the *ENG* subcohort increased with age, noting this could span periods including embolisation (Figure 4c). There was no difference in the relationship between mean PAP and SaO_2_ between the genotypes (Figure 4d).

### Major hemoptysis/hemothorax rates

Hemothorax where bleeding occurs into the pleural space was recorded in 5 cases [0.005, 0.017], and massive hemoptysis affected 13 patients [0.005, 0.017]. For the 18 patients who suffered one of these events, in 8 [0.19, 0.70] this was fatal or required emergency surgical or embolisation intervention. As reported previously for 258/262 [0.97, 1.00] women with PAVMs^15^, most pregnancies did not result in bleeding, but 7/18 massive hemoptysis/hemothorax episodes occurred during pregnancy or the immediate post-partum period.

### Stroke risks

One or more definite clinical strokes affected 246/1149 [0.19, 0.24] patients before review, or during follow-up. A further 53 patients [0.03, 0.06] had an event that could not be confirmed as a stroke (Supplementary Data Item 1).

Examining by stroke type, 125/1,149 [0.09, 0.13] patients had at least one ischemic stroke supported by imaging, and 107/1,149 [0.08, 0.11] had at least one brain abscess (maximum two unrelated events), requiring neurosurgical drainage or causing death. There were lower rates of ischemic strokes and brain abscess in 62 patients with solely ‘tiny/small’ PAVMs, 39 patients where PAVMs were suspected but could not be confirmed (‘possible’ PAVMs), or in 44 patients where a referral diagnosis of PAVMs was subsequently revised to an alternate diagnosis and labelled as ‘PAVM mimics’ (Supplementary Data Item 1). Hemorrhagic strokes affected 23/1,149 [0.01, 0.03], all with confirmed or suspected HHT.

Focusing on stroke-in-young, cerebral hemorrhage tended to occur earlier, with stroke-free survival curves indicating that ischemic stroke became more frequent by ∼24y and brain abscess more frequent by ∼28y (Figure 5a). Over the life-course, Kaplan-Meier estimates were that ∼55% of patients with PAVMs would have an ischemic stroke or brain abscess compared to ∼5% having a hemorrhagic stroke (Figure 5b). Ischemic stroke and brain abscess occurred at substantially lower rates in a comparator cohort of patients with HHT without PAVMs, though ischemic strokes became more frequent after 66y (Figure 5c/d).

**Figure 5:**
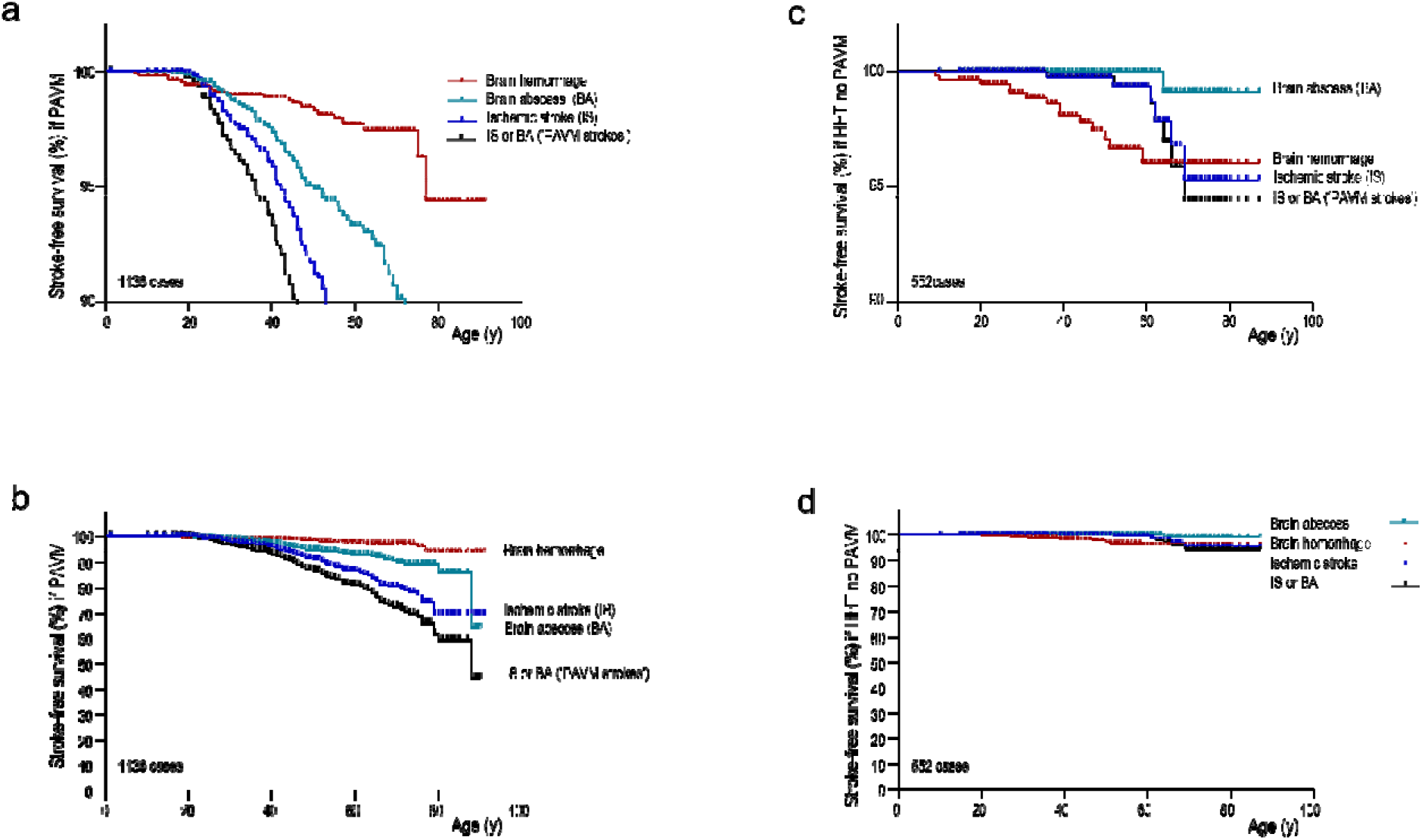
Kaplan-Meier survival curves to first clinical stroke in patients with PAVMs or HHT without PAVMs. Kaplan-Meier survival curves for confirmed stroke types in **a/b)** 1,135 patients with radiologically-confirmed PAVMs (+/-HHT), and **c/d)** 552 HHT patients where PAVMs were excluded by thoracic CT scans. The numbers exclude 29 patients who had a stroke of unknown etiology or a ‘possible stroke’. **a/c)** Time to survival without a clinical stroke of stated type displaying curves to 90% survival to focus on stroke-in-young, and **b/d)** full curves. Note ischemic strokes were confirmed by reported local imaging and prescription of antiplatelet therapies, exclude transient ischemic attacks (TIAs) where symptoms lasted <24h, and exclude cerebral infarcts in the absence of a clinical stroke with duration >24h. Brain abscess was confirmed by imaging reports and need for neurosurgical drainage and 6 weeks intravenous antibiotics; cerebral hemorrhage by imaging and local reports.

### Unmet need for new data

Of 1716 publications after the British Thoracic Society Clinical Statement^2^ retrieved by the PubMed search on 30 April 2026 for ‘pulmonary arteriovenous’, 580 were relevant. Nearly three-quarters of these 580 publications were case reports (415 [72%]). The remaining articles included review articles (60 [10%]); primary data series for interventions or diagnostics (51 [9%] for embolisation/surgery; 23 [4%] for diagnostics), with a small number relevant to medical care^3,16-19^ (Supplementary Data Item 2). The current data were therefore considered a useful addition to the literature, representing the largest PAVM series reported to date.

## DISCUSSION

These data reinforce for modern readerships that PAVMs commonly cause hypoxemia but not hypoxic pulmonary hypertension. PAVMs are seen to be associated generally with low bleeding rates, but hemorrhage can be life-threatening, with major bleeds occurring more often in pregnancy. We also show that PAVMs commonly cause ischemic stroke and brain abscess, that PAVMs account for the majority of strokes in people with HHT, and that PAVM-induced stroke rates exceed those of cerebral hemorrhage. PubMed searches generally retrieving single case reports further emphasise the value of this series of 1149 consecutive cases.

## Data Availability

Data access is via the iCARE Secure Data Environment (https://www.imperial.ac.uk/medicine/research-and-impact/groups/icare/access-to-data/).

## FUNDING/SUPPORT

This research was co-funded by the National Institute for Health Research (NIHR) Imperial Biomedical Research Centre (NIHR203323), HHT charitable donations, and received funding from the NIHR Imperial BRC Digital Health Theme pilot project scheme. The research for PAVM patient data was enabled by the iCARE Secure Data Environment and used the iCARE team and data resources under approved project “Strokes and pulmonary arteriovenous malformations.” iCARE infrastructure support was provided by the NIHR Imperial Biomedical Research Centre (NIHR203323).The sponsors and funding sources played no role in the development of the research or manuscript. The views expressed are those of the authors and not necessarily those of the NHS, the NIHR, or the Department of Health and Social Care.

## ETHICAL APPROVALS

The Imperial iCARE database was given favourable ethics approval by the South West - Central Bristol Research Ethics Committee (reference 21/SW/0120; IRAS project ID 282093). Hammersmith and Queen Charlotte’s & Chelsea Research Ethics Committee (REC ref. 2000/5764) had approved routine data from National Health Service (NHS) clinical care to be used for research and this included generation of a prospective database of patients seen in adult medical PAVM services at Hammersmith Hospital from 1 May 1999-09 April 2026. Within the Imperial iCARE database, a Rare Disease database was constructed to hold the data all patients with known PAVMs reviewed in the Imperial/Hammersmith paediatric and adult PAVM services from 29/05/1984, when patients seen only prior to 1999, or solely in paediatric or embolisation services were added. Data were deidentified by the NIHR Imperial Biomedical Research Centre (BRC) iCARE team before hosting in the iCARE Secure Data Environment. Project approval of the project “Strokes and pulmonary arteriovenous malformations” was by the January 2026 NIHR Imperial BRC Data Access and Prioritisation Committee Meeting.

## ETHICS

Ethical approval for the prospective study was provided by the Hammersmith and Queen Charlotte’s & Chelsea Research Ethics Committee (REC ref. 2000/5764: Hammersmith Hospital patients with pulmonary arteriovenous malformations (PAVMs) and hereditary haemorrhagic telangiectasia) and remains valid. The iCARE research database was given favourable ethics approval by the South West - Central Bristol Research Ethics Committee (reference 21/SW/0120; IRAS project ID 282093).

## DATA AVAILABILITY

Imperial data access is via the iCARE Secure Data Environment (https://www.imperial.ac.uk/medicine/research-and-impact/groups/icare/access-to-data/).

## ACKNOWLEDGMENTS

This research was co-funded by the National Institute for Health Research (NIHR) Imperial Biomedical Research Centre (NIHR203323), HHT charitable donations, and received funding from the NIHR Imperial BRC Digital Health Theme pilot project scheme. The research for PAVM patient data was enabled by the iCARE Secure Data Environment and used the iCARE team and data resources under approved project “Strokes and pulmonary arteriovenous malformations.” iCARE infrastructure support was provided by the NIHR Imperial Biomedical Research Centre (NIHR203323). The views expressed are those of the authors and not necessarily those of the NHS, the NIHR, or the Department of Health and Social Care.

---

**SUPPLEMENTARY DATA ITEM 1:**
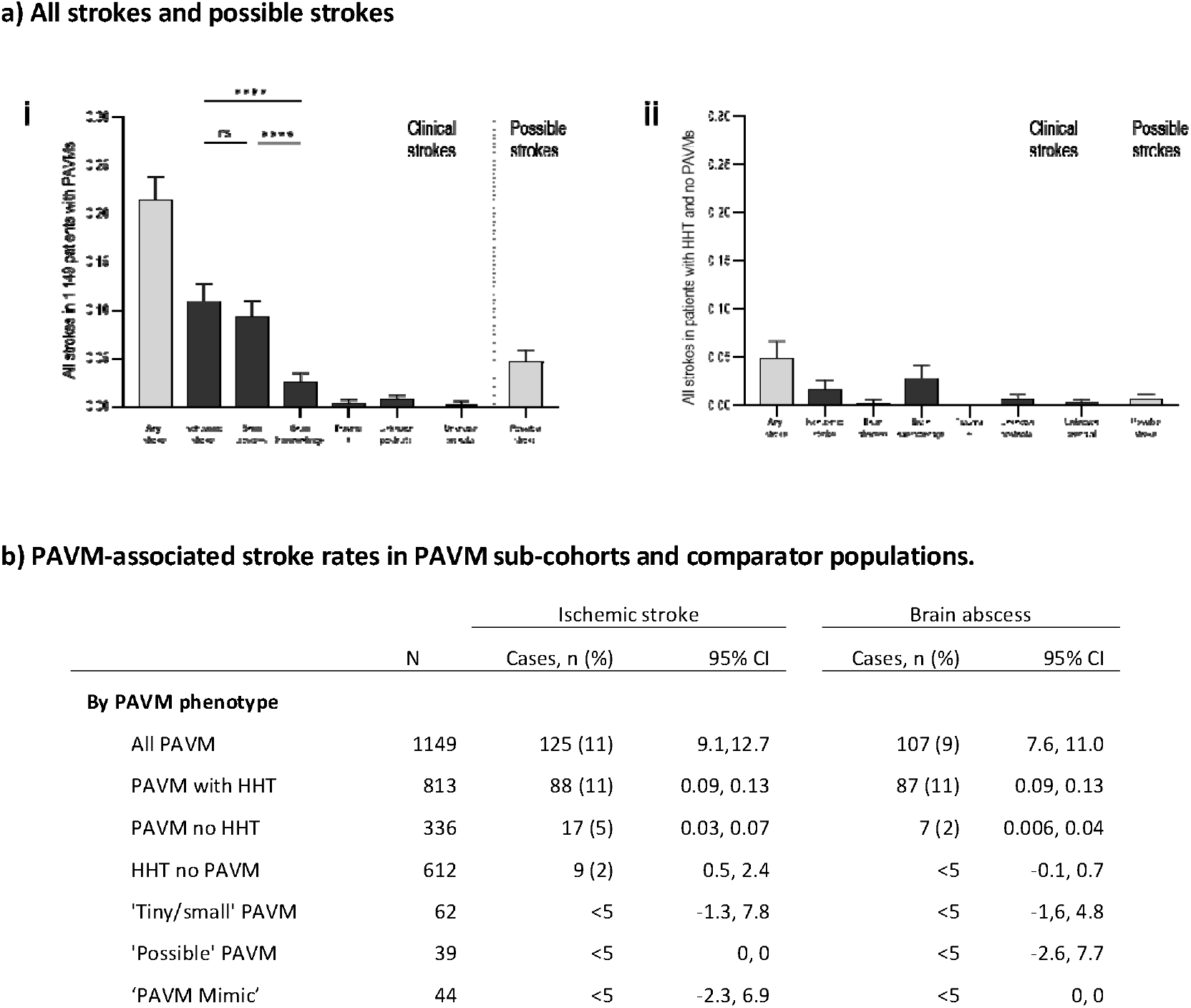
Crude stroke rates in PAVM sub-cohorts. **a)** Crude ischemic stroke and brain abscess rates compared to other types of strokes, brain injury and suspected strokes in **a)** 1,149 patients with radiologically-confirmed PAVMs, including 813 with HHT, and **b)** 567 HHT patients where PAVMs were excluded by thoracic CT scans. Means and 95% confidence intervals of means displayed. P values calculated by Wald test post Kruskal Wallis (**** p<0.001, ns non-significant, p>0.05). Note ischemic strokes were confirmed by reported local imaging and prescription of antiplatelet therapies, exclude transient ischemic attacks (TIAs) where symptoms lasted <24h, and exclude cerebral infarcts in the absence of a clinical stroke with duration >24h. Brain abscess was confirmed by imaging reports and need for neurosurgical drainage and 6 weeks intravenous antibiotics; cerebral hemorrhage by imaging and local reports. ‘Possible strokes’ are where a stroke etiology could be neither excluded nor confirmed for a clinical event of duration >24h. **b)** Rates, percentage and 95% confidence intervals reported for all PAVMs; patients with HHT patients without PAVMs; and patients with small PAVMs (with descriptors of ‘tiny’, ‘very small’ or ‘small’ applied to the only PAVM(s) on the scan); ‘possible’/’likely’ PAVMs, and patients referred to the service with one or more PAVMs that were subsequently excluded due to an alternate diagnosis (‘PAVM mimics).

**SUPPLEMENTARY DATA ITEM 2:**
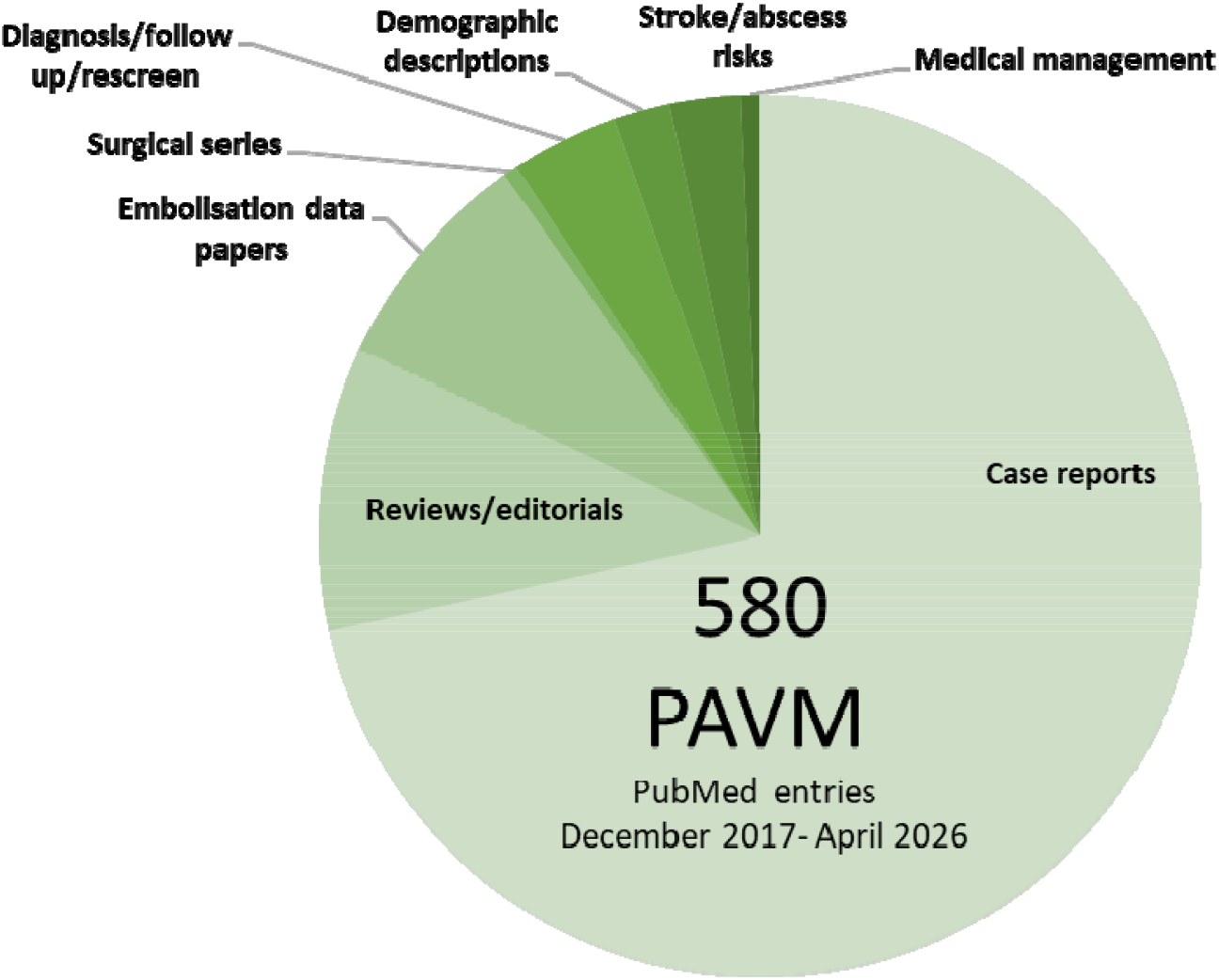
Ten years of PAVM literature 2017-2026. Categories of the 580 PAVM-relevant publications amongst 1716 publications retrieved by a PubMed search on 30 April 2026 for ‘pulmonary arteriovenous’.

## Notes

### Competing Interest Statement

The authors have declared no competing interest.

### Clinical Protocols

https://journals.plos.org/plosone/article?id=10.1371/journal.pone.0088812

### Author Declarations

Ethical approval for the prospective study was provided by the Hammersmith and Queen Charlottes & Chelsea Research Ethics Committee (REC ref. 2000/5764: Hammersmith Hospital patients with pulmonary arteriovenous malformations (PAVMs) and hereditary haemorrhagic telangiectasia) and remains valid. The research database was given favourable ethics approval by the South West - Central Bristol Research Ethics Committee (reference 21/SW/0120; IRAS project ID 282093).

### Summary of Updates

HHT gene and time-series comparisons have been used to update the physiological data presented . Kaplan-Meier survival curves have been added for stroke risks.

## REFERENCES

1 Shovlin CL. Pulmonary arteriovenous malformations. Am J Respir Crit Care Med. 2014 Dec 1;190(11):1217–28.

2 The British Thoracic Society Clinical Statement on pulmonary arteriovenous malformations. Thorax. 2017 Dec;72(12):1154–1163

3 Topiwala KK, Patel SD, Pervez M, et al. Ischemic stroke in patients with pulmonary arteriovenous fistulas. Stroke. 2021 Jul;52(7):e311–e315.

4 Boother EJ, Brownlow S, Tighe HC et al. Cerebral abscess associated with odontogenic bacteremias, hypoxemia, and iron loading in immunocompetent patients with right-to-left shunting through pulmonary arteriovenous malformations. Clin Infect Dis. 2017 Aug 15;65(4):595–603

5 Nakayama M, Nawa T, Chonan T, et al. Prevalence of pulmonary arteriovenous malformations as estimated by low-dose thoracic CT screening. Intern Med. 2012;51(13):1677–81

6 Hermann R, Shovlin CL, Kasthuri RS, et al. Hereditary haemorrhagic telangiectasia. Nat Rev Dis Primers. 2025 Jan 9;11(1):1.

7 Müller-Hülsbeck S, Marques L, Maleux G, Osuga K, Pelage JP, Wohlgemuth WA, Andersen PE. CIRSE Standards of Practice on diagnosis and treatment of pulmonary arteriovenous malformations. Cardiovasc Intervent Radiol. 2020 Mar;43(3):353–361

8 Alsafi A, Shovlin CL. Orphanet Definition of Pulmonary Arteriovenous malformation. 2021, available at https://www.orpha.net/en/disease/detail/2038

9 Shovlin CL, Chamali B, Santhirapala V, Livesey JA, Angus G, Manning R, Laffan MA, Meek J, Tighe HC, Jackson JE. Ischaemic strokes in patients with pulmonary arteriovenous malformations and hereditary hemorrhagic telangiectasia: associations with iron deficiency and platelets. PLoS One. 2014 Feb 19;9(2):e88812.

10 Shovlin CL, Tighe HC, Davies RJ, Gibbs JS, Jackson JE. Embolisation of pulmonary arteriovenous malformations: no consistent effect on pulmonary artery pressure. Eur Respir J. 2008 Jul;32(1):162–9

11 Shovlin CL, Gibbs JS, Jackson JE. Management of pulmonary arteriovenous malformations in pulmonary hypertensive patients: a pressure to embolise? Eur Respir Rev. 2009 Mar;18(111):4–6.

12 Shovlin CL, Guttmacher AE, Buscarini E, et al. Diagnostic criteria for hereditary hemorrhagic telangiectasia (Rendu-Osler-Weber syndrome). Am J Med Genet. 2000 Mar 6;91(1):66–7.

13 Kovacs G, Berghold A, Scheidl S, Olschewski H. Pulmonary arterial pressure during rest and exercise in healthy subjects: a systematic review. Eur Respir J. 2009 Oct;34(4):888–94.

14 Jutant EM, Grynblat J, Pyrrait M, et al. Pulmonary hypertension associated with hereditary haemorrhagic telangiectasia: from genetics to clinical management. Eur Respir J. 2026 Apr 16;67(4):2502322

15 Shovlin CL, Sodhi V, McCarthy A, Lasjaunias P, Jackson JE, Sheppard MN. Estimates of maternal risks of pregnancy for women with hereditary haemorrhagic telangiectasia (Osler-Weber-Rendu syndrome): suggested approach for obstetric services. BJOG. 2008 Aug;115(9):1108–15

16 Gawecki F, Strangeways T, Amin A, Perks J, McKernan H, Thurainatnam S, Rizvi A, Jackson JE, Santhirapala V, Myers J, Brown J, Howard LSGE, Tighe HC, Shovlin CL. Exercise capacity reflects airflow limitation rather than hypoxaemia in patients with pulmonary arteriovenous malformations. QJM. 2019 May 1;112(5):335–342.

17 Alsafi A, Jackson JE, Fatania G, Patel MC, Glover A, Shovlin CL. Patients with in-situ metallic coils and Amplatzer vascular plugs used to treat pulmonary arteriovenous malformations since 1984 can safely undergo magnetic resonance imaging. Br J Radiol. 2019 ;92(1098):20180752

18 Delagrange L, Dupuis O, Fargeton AE, Bernard L, Decullier E, Dupuis-Girod S. Obstetrical and neonatal complications in hereditary haemorrhagic telangiectasia: A retrospective study. BJOG. 2023 Feb;130(3):303–311

19 Das H, Ironton HR, Coote NM, Cabantug JA, Ranger J, Alsafi A, Shovlin CL. Adolescent exercise capacity predicts higher exercise tolerance later in life when compounded by anaemia in patients with pulmonary arteriovenous malformations. Thorax. 2025 Sep 15;80(10):756–760

